# Severity Assessment of COVID-19 based on Clinical and Imaging Data

**DOI:** 10.1101/2020.08.12.20173872

**Authors:** Juan C. Quiroz, You-Zhen Feng, Zhong-Yuan Cheng, Dana Rezazadegan, Ping-Kang Chen, Qi-Ting Lin, Long Qian, Xiao-Fang Liu, Shlomo Berkovsky, Enrico Coiera, Lei Song, Xiao-Ming Qiu, Sidong Liu, Xiang-Ran Cai

## Abstract

**Objectives:** This study aims to develop a machine learning approach for automated severity assessment of COVID-19 patients based on clinical and imaging data.

**Materials and Methods:** Clinical data—demographics, signs, symptoms, comorbidities and blood test results—and chest CT scans of 346 patients from two hospitals in the Hubei province, China, were used to develop machine learning models for automated severity assessment of diagnosed COVID-19 cases. We compared the predictive power of clinical and imaging data by testing multiple machine learning models, and further explored the use of four oversampling methods to address the imbalance distribution issue. Features with the highest predictive power were identified using the SHAP framework.

**Results:** Targeting differentiation between mild and severe cases, logistic regression models achieved the best performance on clinical features (AUC:0.848, sensitivity:0.455, specificity:0.906), imaging features (AUC:0.926, sensitivity:0.818, specificity:0.901) and the combined features (AUC:0.950, sensitivity:0.764, specificity:0.919). The SMOTE oversampling method further improved the performance of the combined features to AUC of 0.960 (sensitivity:0.845, specificity:0.929).

**Discussion:** Imaging features had the strongest impact on the model output, while a combination of clinical and imaging features yielded the best performance overall. The identified predictive features were consistent with findings from previous studies. Oversampling yielded mixed results, although it achieved the best performance in our study.

**Conclusions:** This study indicates that clinical and imaging features can be used for automated severity assessment of COVID-19 patients and have the potential to assist with triaging COVID-19 patients and prioritizing care for patients at higher risk of severe cases.

## BACKGROUND AND SIGNIFICANCE

Coronavirus disease 2019 (COVID-19) has overwhelmed health systems worldwide. [1,2] As of July 5, 2020, more than 11 million cases have been confirmed worldwide, with 528,953 global deaths.[3] Given the various complications associated with COVID-19,[4-6] methods that facilitate triage of COVID-19 can help prioritize care for those who are likely to experience severe or critical cases. COVID-19 illness severity can be defined as four groups: (1) mild, (2) ordinary, (3) severe, and (4) critical.[7] Severe and critical cases require intensive care and more healthcare resources. A high rate of false positive severe or critical cases could overwhelm healthcare resources (i.e., ICU beds). Equally important, delays in identifying severe or critical cases would lead to delayed treatment of patients at a higher risk of mortality. It is, therefore, important to identify severe cases as early as possible, so that resources can be mobilized and treatment can be escalated.

Chest CT scans have been found to provide important diagnostic and prognostic information,[8,9] and consequently, they have been the focus of numerous recent studies using machine learning techniques for prediction tasks related to the COVID-19 pandemic.[10-22] Studies have looked at mortality predictions,[10] diagnosis (detecting COVID cases and differentiating from other pulmonary diseases or no disease),[11,15-19] and severity assessment and disease progression.[20-22] The majority of current approaches have used deep learning and imaging features from CT-scans[11,12,15-19] and X-rays,[13,14,22] with popular architectures including ResNet,[11,16,18] U-Net,[15,21] Inception,[19] Darknet,[13] and other convolutional neural networks.[14,22] More details can be found in recent review papers[1,23-25].

Automatic assessment of chest CT scans to predict COVID-19 severity is of a great clinical importance, but has only been the focus of few studies.[20-22] Automated assessment of chest CT scans can substantially reduce the image reading time for radiologists, provide quantitative information that can be compared across patients and time-points, with clinical applications in detection and diagnosis, progression tracking and prognosis.[9] While CT scans are an important diagnostic tool, prior work has also shown that clinical data, such as symptoms, comorbidities and laboratory findings, differed for COVID-19 patients who were admitted to intensive care units (ICU) vs non-ICU patients,[26] and were predictive of the mortality risk.[10] One study compared the imaging data and clinical data of 81 confirmed COVID-19 patients, suggesting that the combination of imaging features with clinical and laboratory findings could facilitate early diagnosis of COVID-19.[27]

In this study, we used patient clinical data and imaging data to predict COVID-19 case severity. We consider this as a binary classification task, predicting whether a diagnosed patient is likely to develop a mild or a severe case of COVID. The contributions of this work are three-fold. First, we compared the predictive power of clinical and image data for severity assessment, by testing three machine learning models: logistic regression (LR),[28] gradient boosted trees (XGB),[29] and neural network (NN).[30] Secondly, due to the cohort data being highly imbalanced, with the majority of cases being mild/ordinary, we explored the use of four oversampling methods to address the imbalance distribution issue.[31-34] Third, we interpreted the importance of features using the SHAP (SHapley Additive exPlanations) framework and identified the features with the highest predictive power.[35] The evaluated predictive models yielded high accuracy and identified predictive imaging and clinical features consistent with prior findings.

## MATERIALS AND METHODS

### Study Design and Participants

This is a retrospective study carried out with data collected by two hospitals in the Hubei province, China. The study cohort consisted of patients who had COVID-19 diagnosis confirmed by reverse transcription PCR (RT-PCR). A total of 346 patients from two hospitals were retrospectively enrolled, including 230 patients from Huang Shi Central Hospital (HSCH) and 116 from Xiang Yang Central Hospital (XYCH). These patients were admitted to hospital between 11-01-2020 and 23-02-2020, and all underwent chest CT scans at initial hospitalization. All the participants provided written consent. This study was approved by the Institutional Review Board of both hospitals. **Table 1** shows the demographics of the two cohorts of patients.

**Table 1.** Demographics of the two cohorts of patients.

| Category | HSCH | XYCH | Total |
| --- | --- | --- | --- |
| Mild | 7 | 1 | 8 |
| Ordinary | 212 | 104 | 316 |
| Severe | 7 | 6 | 13 |
| Critical | 4 | 5 | 9 |
| Total patients | 230 | 116 | 346 |
| Age (mean±SD) | 49.0±14.4 | 47.5±17.2 | 48.5±15.4 |
| Gender (female/male) | 120/110 | 57/59 | 177/169 |

### Imaging and Clinical Data

Chest CT scans were collected from the patients at initial hospitalization. All CT scans were pre-processed with intensity normalization, contrast limited adaptive histogram equalization, and gamma adjustment, using the same pre-processing pipeline as in our previous study.[36] We performed lung segmentation on the CT slices using an established model – R231CovidWeb,[37] trained on a large and diverse dataset of non-COVID-19 CT scans and further fine-tuned with an additional COVID-19 dataset.[38] The CT slices with less than 3mm^2^ lung tissue were removed from the datasets since they bear little or no information of the lung. The lesions were segmented using EfficientNetB7 U-Net,[20] also trained using a public COVID-19 dataset.[38] The model produced four types of lesions, including ground-glass opacities, consolidations, pleural effusions, and other abnormalities. The volume of each lesion type and the total lesion volume were calculated from the segmentation maps as the imaging features, which were further normalized by the lung volume. **Figure 1** shows examples of the lung and lesion segmentation results of a mild case and a severe case. The upper row presents the 3D models of the lung and lesions, reconstructed using 3D Slicer (v4.6.2),[39] and the lower row presents the axial CT slices with the lung and lesion (green: ground-glass opacities; yellow: consolidation; brown: pleural effusion) boundaries overlaid on the CT slices.

**Figure 1.**
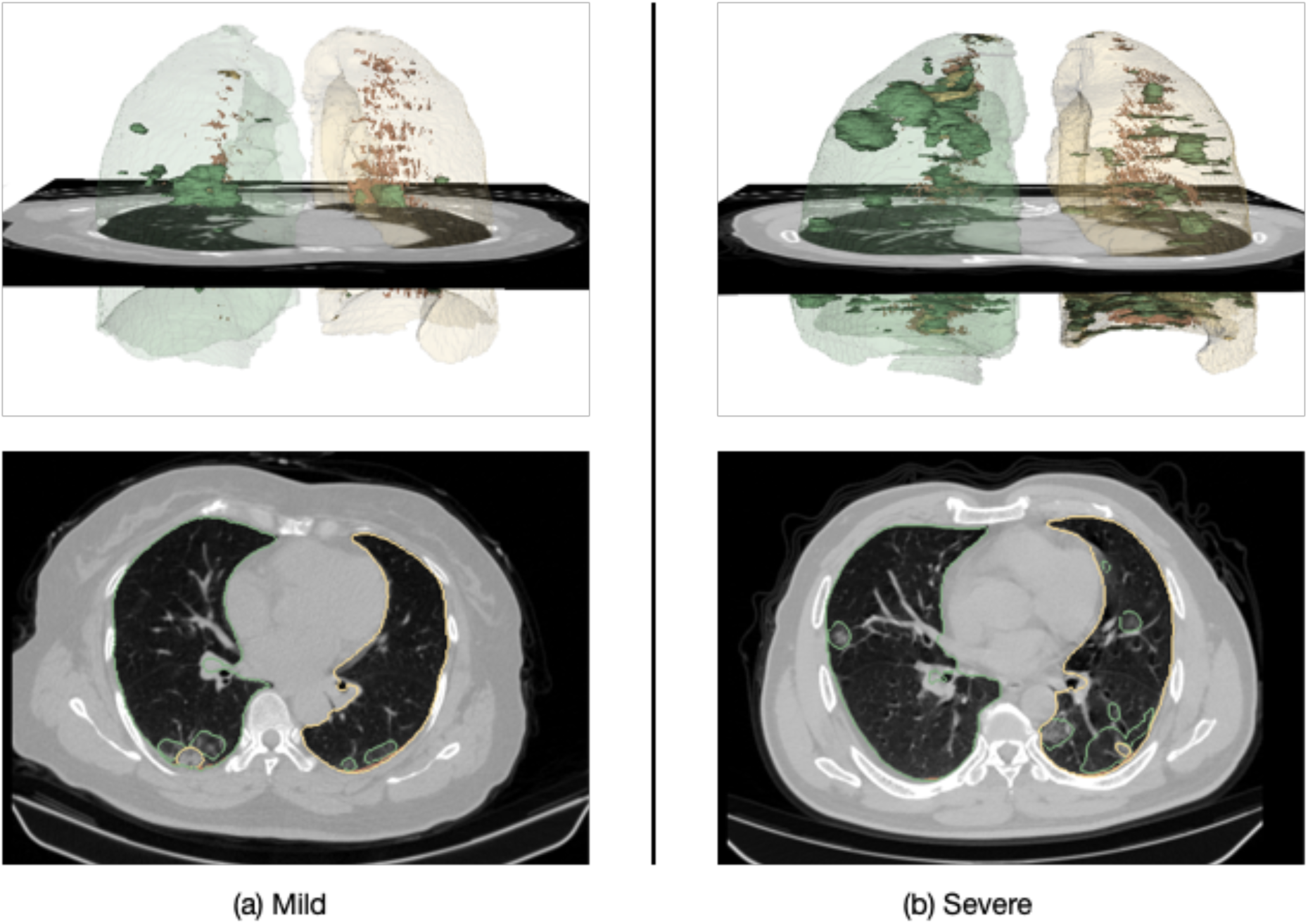
Examples of the CT scans and the lung and lesion models of (a) mild case and (b) a severe case.

Clinical data collected from the patients included demographics, signs, symptoms, comorbidities and 18 laboratory test results: white blood cell count (× 10^9^/L), neutrophil count (× 10^9^/L), lymphocyte count (× 10^9^/L), haemoglobin (× 10^9^/L), platelet (× 10^9^/L), prothrombin time (s), activated partial thromboplastin time (s), D-dimer (mg/L), C-reactive protein (mg/L), albumin (g/L), alanine aminotransferase (U/L), aspartate aminotransferase (U/L), total bilirubin (mmol/L), potassium (mmol/L), sodium (mmol/L), creatinine (p.mol/L), creatine kinase (U/L), and lactate dehydrogenase (U/L).

All the features were either continuous or binary—all binary features belong to signs, symptoms and comorbidities. Continuous features were standardized to be centred around 0 with a standard deviation of 1. **Figure 2** shows the structure and dimensions of the features used in this study.

**Figure 2.**
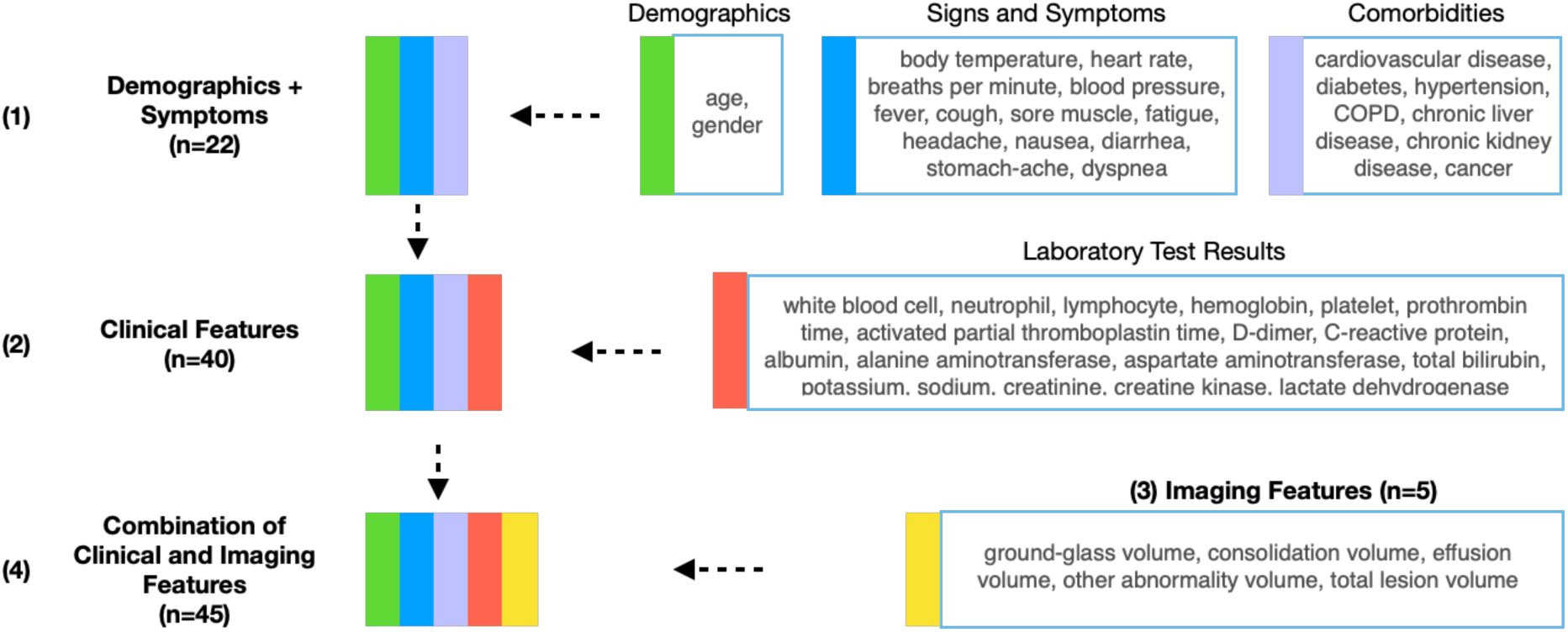
Structure and dimensions of the feature sets.

These features were grouped into four feature sets: (1) demographic and symptoms (a subset of the available clinical features), (2) clinical features (demographic, signs and symptoms, and laboratory test results), (3) imaging features extracted from the CT chest scans using deep learning, and (4) combination of clinical and imaging features.

### Severity Assessment Models

Three models were trained and compared to predict case severity: logistic regression (with Scikit-learn),[40] gradient boosted trees (with XGBoost),[29] and a neural network (with Fastai).[41] We used the HSCH data (230 samples) for training and validation using 5-fold repeated stratified cross-validation. The XYCH data (116 samples) was withheld for testing. We report results for the test set using AUC and F1 scores averaged over the independent runs.

Hyperparameter exploration and tuning were done using the train/validation set. Random search was used to tune the hyperparameters of LR and XGB. For NN, we used a four-layer, fully-connected architecture, with the first hidden layer having 200 nodes and the second hidden layer having 100 nodes. The learning rate (0.01) was determined using the learning rate finder.[42] All other parameters of NN were set to default values. We explored a different number of nodes in the first and second hidden layers, with 200×100 yielding the best results in the validation set. Out of the 346 patients, 167 (48%) had at least one missing feature (5.7 on average, mostly in the laboratory test results category). Missing feature values were imputed with the mean for each feature.

### Oversampling

The majority of cases in our dataset were mild/ordinary cases and the minority were severe/critical cases. The imbalance ratio for the entire dataset was 0.07, for the training/validation set – 0.05, and for the testing set – 0.10. We tested four oversampling methods to increase the ratio of the minority class: Synthetic Minority over-Sampling Technique (SMOTE),[31] ADAptive SYNthetic sampling approach (ADASYN),[32] geometric SMOTE,[33] and a conditional generative adversarial network model for tabular data (CTGAN).[34] For all these methods, we oversampled the training set, trained a model on the oversampled data, and reported results on the same test set. We fixed the resampling ratio of all methods to 0.3 (bringing imbalance ratio to 0.3). When using CTGAN for oversampling, we fitted the CTGAN model with the training set, sampled to generate synthetic data, keeping only synthetic data for the minority class (severe/critical), repeating until the ratio of the minority to the majority class reached 0.3.

## RESULTS

### Patient Characteristics

**Table 2** presents the patients’ characteristics. The differences between the mild/ordinary and severe/critical groups were calculated with the Mann-Whitney U test and Fisher’s exact test. In the full cohort, the median age was 49 (IQR 38-59). The median age for patients with mild/ordinary cases was 48.5 (IQR 37 – 57.3) and for the severe/critical cases it was 63 (IQR 52.5 – 69.5). There are statistically significant differences between patients with severe/critical and mild/normal infections with respect to age (P < 0.001) and comorbidities of cardiovascular disease (P=0.002), hypertension (P=0.002), diabetes (P=0.01), and cancer (P=0.01). Out of all the signs and symptoms, raised respiration rate (P=0.002) and dyspnea (P<0.001) were more common in patients with severe/critical cases of COVID-19.

**Table 2.**
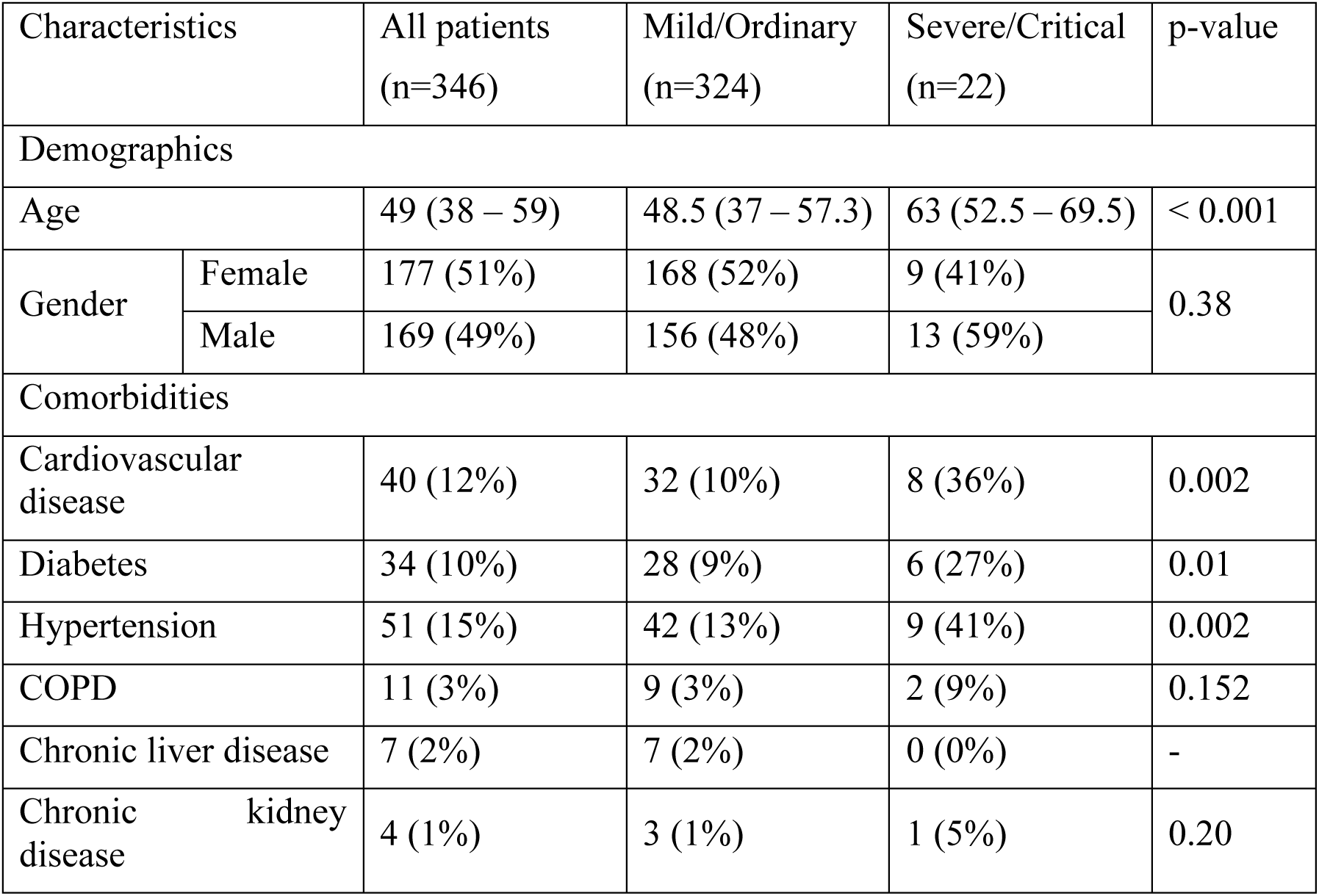
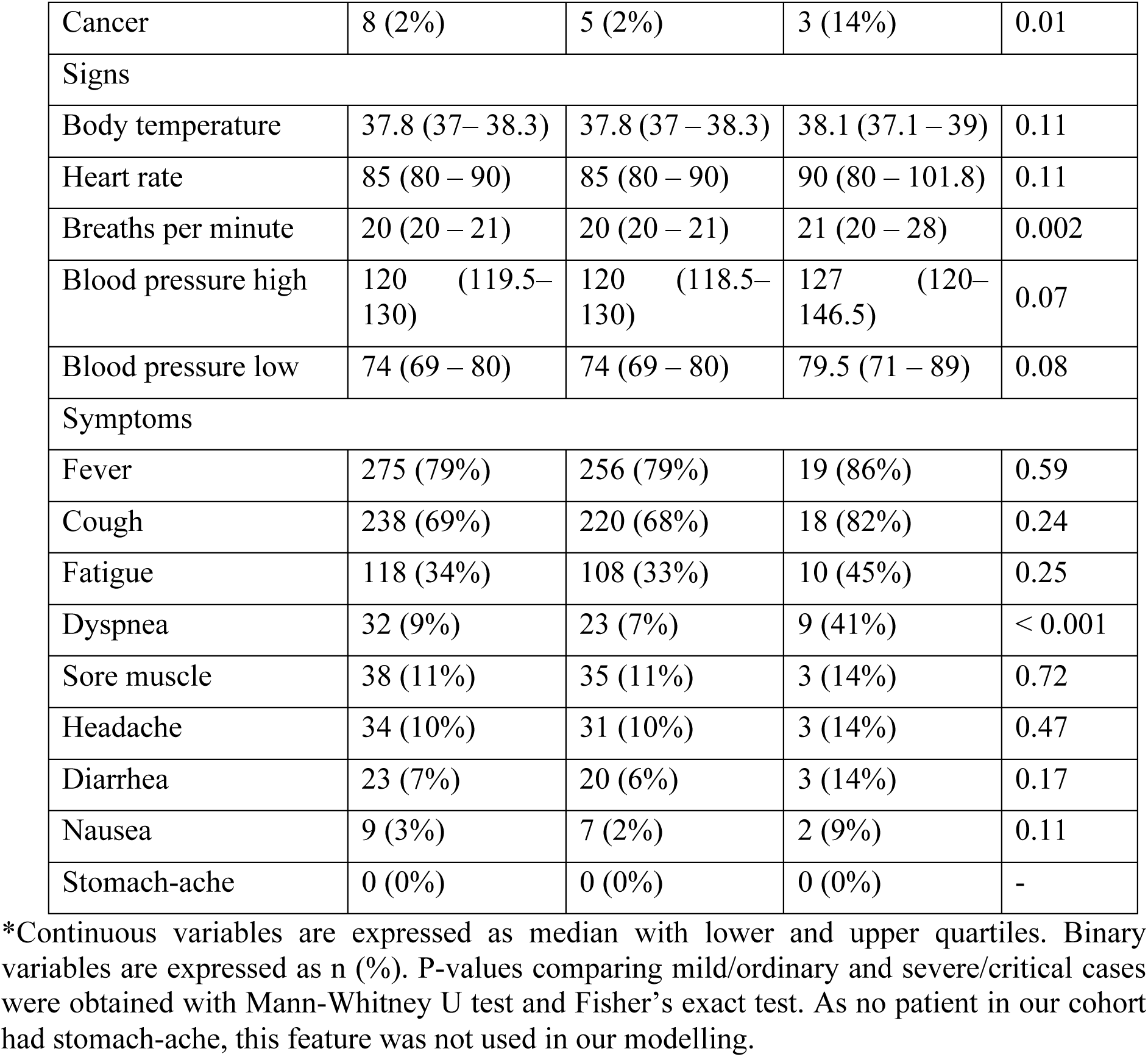
Demographics and baseline characteristics of patients with confirmed COVID-19. Patients with higher severity of COVID-19 were more likely to have cardiovascular disease and experience shortness of breath as a symptom.

### Prediction of COVID-19 Severity at Baseline

The data from HSCH (230 patients) was used for training and validation, and the data from XYCH (116 patients) was used as the independent test set. We compared model performance using four feature sets: (1) demographics and symptoms, (2) clinical features, (3) imaging features and (4) combination of clinical and imaging features, as shown in **Figure 2**. The optimal classification threshold for the sensitivity, specificity and F1 score was identified using Youden’s index.[43] **Table 3** shows the severity assessment performance of an LR model, an XGB model, and a 4-layer fully connected NN model. Overall, LR models outperformed the other evaluated models, achieving the highest AUC, F1 score and sensitivity for all four feature sets. While imaging features yielded substantially better results than clinical features, the combination of clinical and imaging features benefited LR only. Hence, LR yielded the best performance (AUC = 0.950, F1 Score = 0.604, sensitivity = 0.764, specificity = 0.919) using the combination of clinical and imaging features.

**Table 3.**
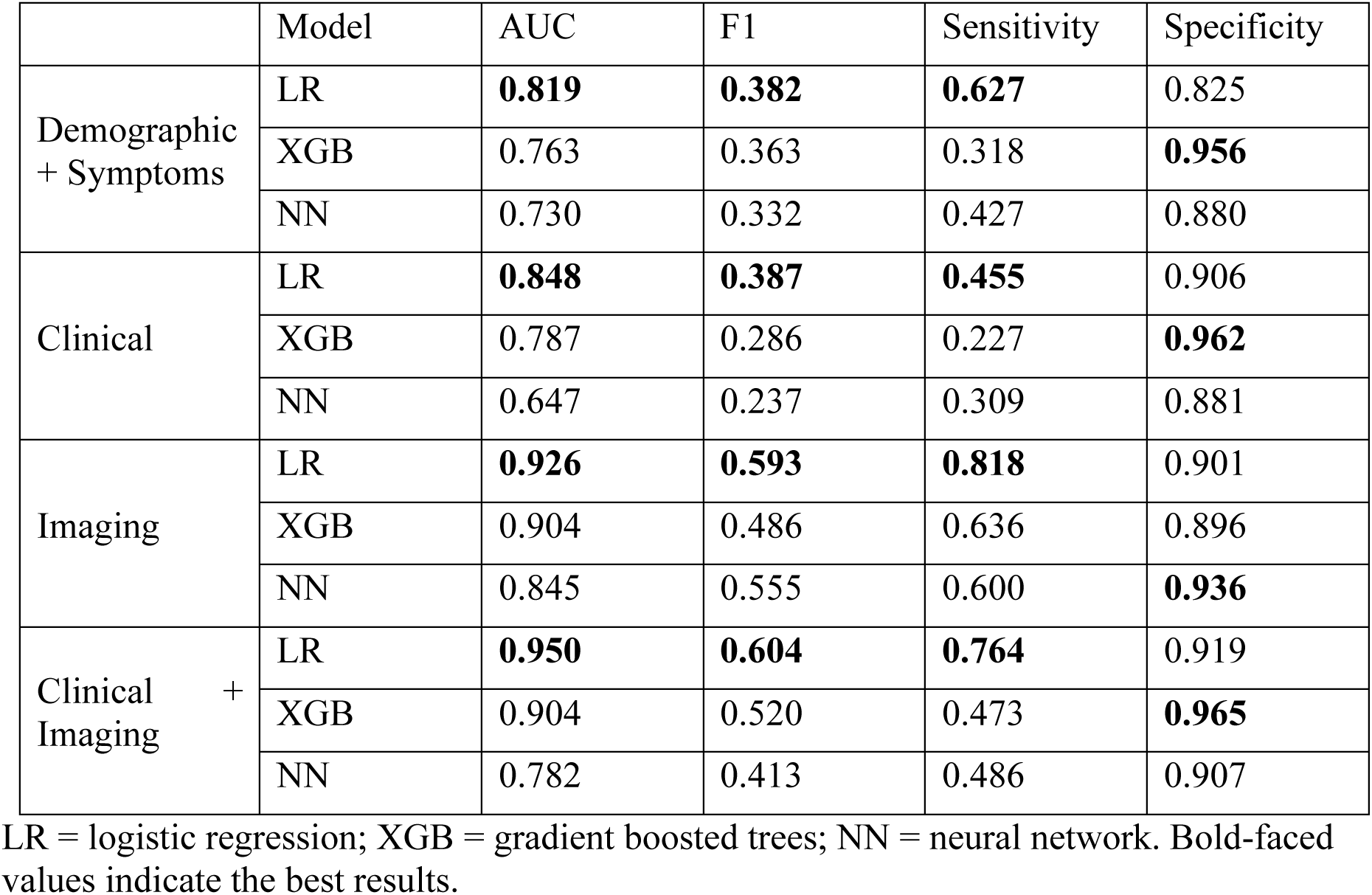
Results from using different feature sets.

### Prediction at Baseline Severity with Oversampling

Since the cohort was highly imbalanced, with the majority of cases being mild/ordinary (imbalance ratio of 0.07), we applied four oversampling methods to increase the ratio of severe/critical cases: SMOTE,[31] ADASYN,[32] geometric SMOTE,[33] and CTGAN.[34] **Figure 3** shows the differences in AUC values and F1 scores resulting from the use of oversampling, with negative values indicating a decrease in AUC or F1 score and positive values indicating the opposite. Oversampling resulted in greater improvements in F1 score compared to AUC. The greatest improvement in F1 (0.09) is observed for the clinical features (Clinical) with XGB and SMOTE method (XGB-smo); however, the AUC dropped by 0.08 with the same method. Considering both AUC and F1 score at the same time, the combination of clinical and imaging features (Clinical + Imaging) benefited the most from oversampling. Specifically, the AUC and F1 score for Clinical + Imaging features were increased by 0.01 and 0.06, respectively, using LR with SMOTE (LR-smo).

**Figure 3.**
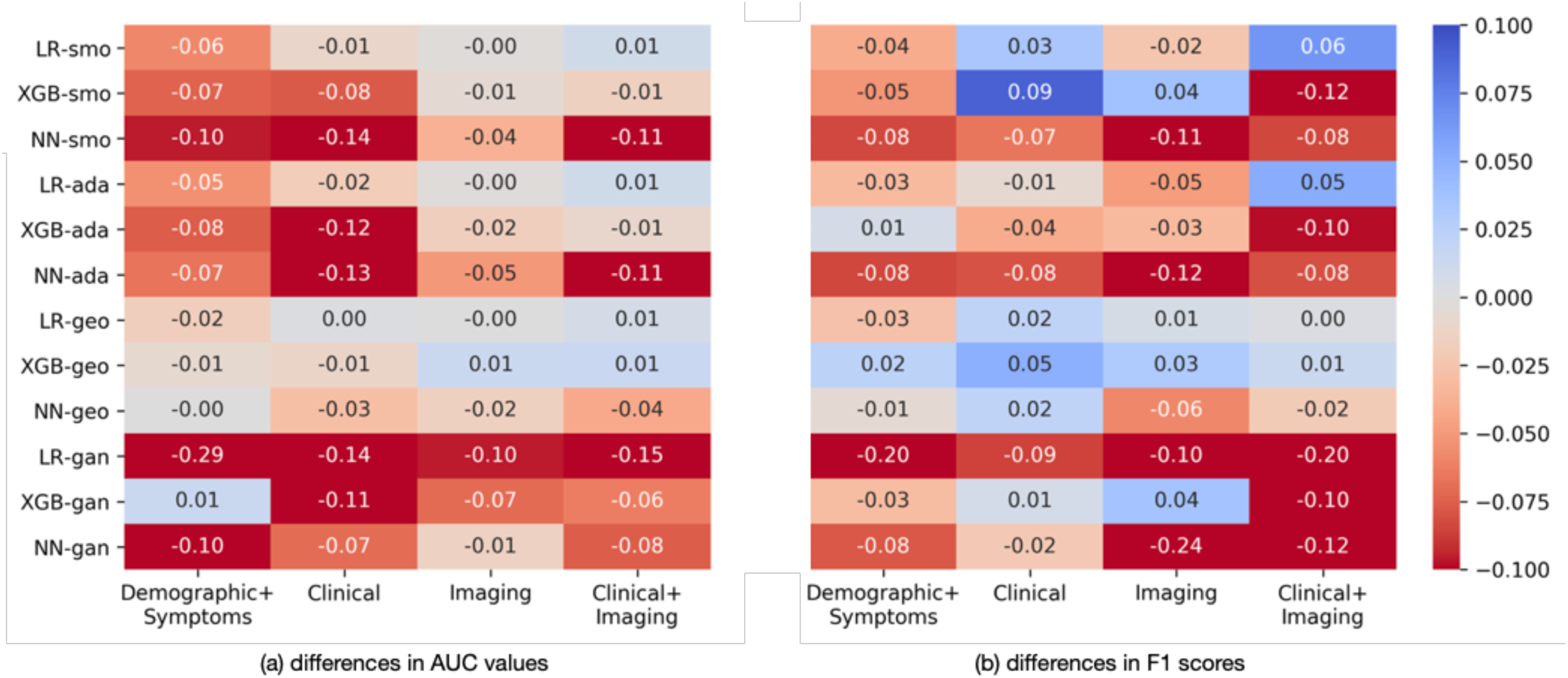
Differences in (a) AUC values and (b) F1 scores with oversampling compared to those without oversampling. Positive values (blue) indicate oversampling resulting in higher values, negative values (red) indicate oversampling resulting in lower values. smo = SMOTE; ada = ADASYN; geo = geometric SMOTE; gan = CTGAN; LR = logistic regression, NN = neural network, XGB = gradient boosted trees.

**Table 4** presents the best results of the evaluated models using various feature sets after oversampling. Oversampling did not improve LR’s performance on Demographics + Symptoms features, but SMOTE and geometric SMOTE resulted in increased F1 scores using Clinical features and Imaging features, respectively. Notably, the best performing in **Table 3** LR model using a combination of clinical and imaging features further improved to AUC of 0.960 (vs. 0.950), F1 score of 0.668 (vs. 0.604), sensitivity of 0.845 (vs. 0.764) and specificity of 0.929 (vs. 0.919), after oversampling with SMOTE.

**Table 4.** The best results from using different feature sets after oversampling.

|  | Model | AUC | F1 | Sensitivity | Specificity |
| --- | --- | --- | --- | --- | --- |
| Demographic + Symptoms | LR* | 0.819 | 0.382 | 0.627 | 0.825 |
| Clinical | LR – smo | 0.837 | 0.421 ↑ | 0.518 ↑ | 0.902 |
| Imaging | LR – geo | 0.926 | 0.599 ↑ | 0.818 | 0.904 ↑ |
| Clinical + Imaging | LR – smo | 0.960 ↑ | 0.668 ↑ | 0.845 ↑ | 0.929 ↑ |

smo = SMOTE; geo = geometric SMOTE; LR = logistic regression; \*no improvement after oversampling; ↑improved performance after oversampling.

### Model Interpretation

We used the SHAP (SHapley Additive explanations) framework[35] to interpret the output of the best performing LR model with SMOTE oversampling. This framework calculates the importance of a feature by comparing model predictions with and without the feature. **Figure 4** illustrates a SHAP plot summarizing how the values of each feature impact the model output of the LR model using all features (clinical and imaging features), with features sorted from most important to least important. **Figure 4(a)** shows feature importance scores sorted by the average impact on the model output, and **Figure 4(b)** presents the SHAP values of individual feature instances. Four imaging features, including consolidation volume (consolidation_val), total lesion volume (lesion_vol), ground-glass volume (groundglass_vol), and volume of other abnormalities (other_vol), are among the top six features with their high values resulting in the model being more likely to predict a severe/critical case of COVID-19. Low albumin, high counts of C-reactive protein, high counts of leukocytes, and low values of lactate dehydrogenase make the model more likely to predict a case severity of critical/severe. Older age and male gender also made the model more likely to predict severe/critical cases.

**Figure 4.**
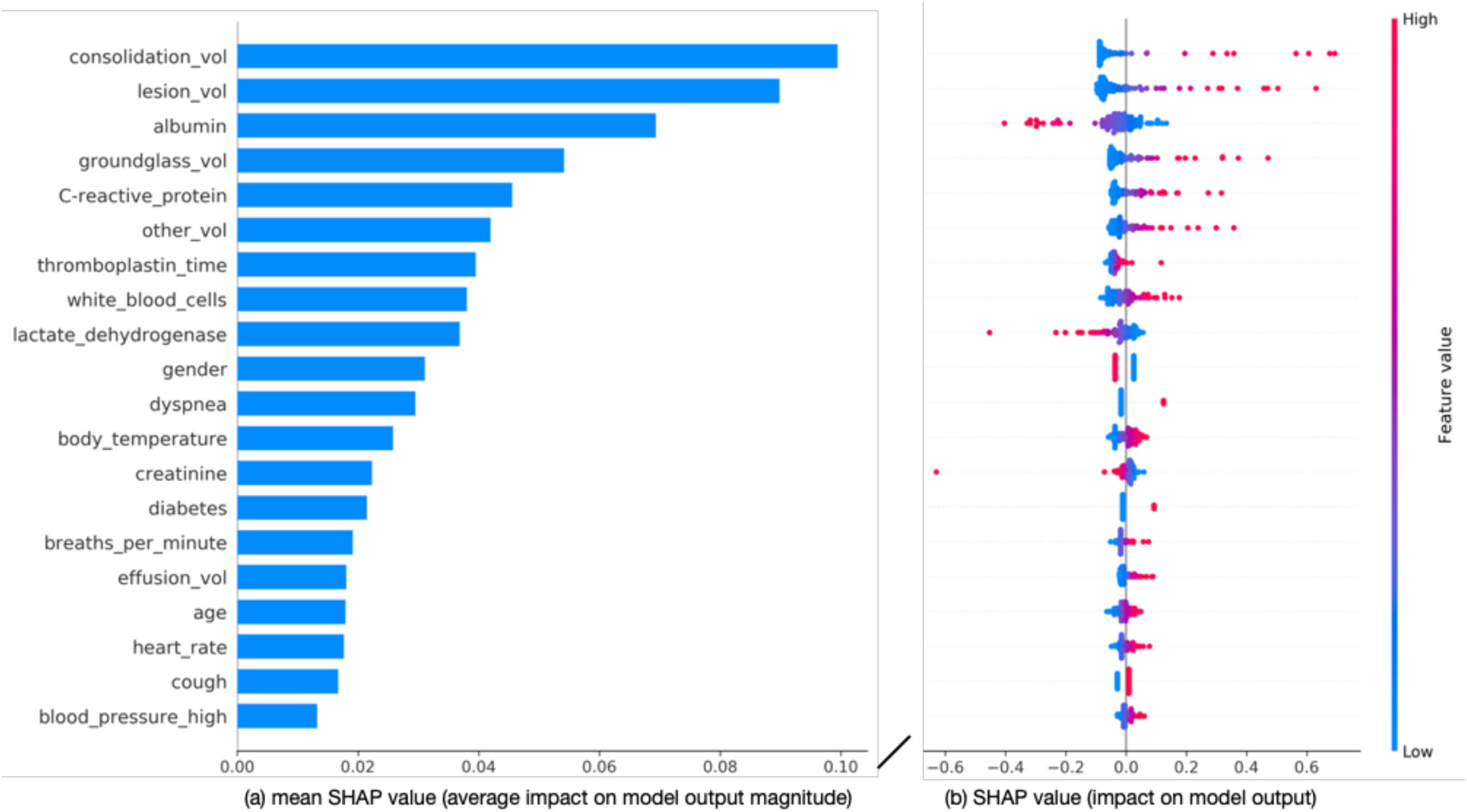
(a) Feature importance, evaluated using the mean SHAP values, in the LR model using all features. (b) SHAP plot for the LR model using all features. Each point represents a feature instance, and the color indicates the feature value (red for high and blue for low). Negative SHAP values indicate feature instances contributing to a model output of a mild/ordinary case of COVID-19, whereas positive SHAP values indicate features contributing to a model output of a severe/critical case.

## DISCUSSION

### Main Findings

In our cohort, fever, cough, and fatigue were the most common symptoms in patients with COVID-19, consistent with other studies of COVID-19 patients.[27] Severe cases manifested a statistically higher incidence of dyspnea and raised respiratory rate. Some symptoms such as sore muscle, headache, diarrhea, and nausea were present in 3-11% of patients and were not statistically different between mild and severe cases. Severe cases of COVID-19 tended to be of older age and had medical comorbidities (cardiovascular disease, diabetes, hypertension, cancer), in similar to prior studies.[1,4,6,27] There was no difference between males and females in our cohort, although the model did rely on gender for increasing the likelihood of predicting a severe/critical case.

A combination of clinical and imaging features yielded the best performance. Imaging features had the strongest impact on the model output, with high values of consolidation volume, lesion volume, ground-glass volume, and other volume making the model more likely to predict a severe case of COVID-19. Ground-glass opacity has also been found to be an important feature in prior work.[18] Inclusion of clinical features further improved the accuracy of severity assessments, with findings such as albumin, C-reactive protein, thromboplastin time, white blood cell counts, and lactate dehydrogenase being amongst the most informative features. The identification of lactate dehydrogenase, white blood cell counts, and C-reactive protein as informative features is consistent with findings from one prior study that also used laboratory findings for COVID-19 mortality prediction.[10] C-reactive protein was also associated with a significant risk of critical illness in a study of 5,279 laboratory confirmed COVID-19 patients.[6] Symptoms and patient characteristics such as gender, dyspnea, body temperature, diabetes, and breaths per minute were also relied on by the model for differentiating between mild and severe cases. Clinical features alone (demographics, signs, symptoms, and laboratory results), resulted in low sensitivity. Relying only on clinical features, thus, poses the risk of predicting mild/ordinary severity for patients who will develop a critical/severe case of COVID-19.

Oversampling yielded mixed results, although it resulted in the best model performance in our study. We note that the best model without oversampling (LR) also yielded strong results (AUC: 0.950, F1: 0.604, sensitivity: 0.764, specificity: 0.919), and the SMOTE oversampling method improved the performance further (AUC: 0.960, F1: 0.668, sensitivity: 0.845, specificity: 0.929). Given the propensity of imbalanced data in healthcare,[44-47] our results suggest the need for further analysis of oversampling methods for medical datasets. Self-supervision,[48,49] may also help in improving performance on imbalanced medical datasets; in particular, future work should evaluate the impact of self-supervision on tabular medical data.

### Clinical Implications

The rapid spread of COVID-19 has put a strain on healthcare systems, necessitating efficient disease severity assessment of COVID-19 patients. Results from this study indicate that clinical and imaging features can inform automated severity assessment of COVID-19. While our work would benefit from a larger dataset, our current results are encouraging given that the models were trained on data from one hospital only and tested on an independent dataset from another hospital, demonstrating nevertheless strong predictive accuracy.

The proposed methods and models would be useful in several clinical scenarios. First, the proposed models are fully automated and can expedite the assessment process, saving time in reading the CT scans or evaluating patients using a scoring system. They can be of use in hospitals that are overwhelmed by a high volume of patients during the outbreak by identifying severe cases as early as possible, so that treatment can be escalated. Our models, with a higher specificity and relatively lower sensitivity, would best be used in combination with a model with higher sensitivity in diagnostic situations, i.e., a high sensitivity model can identify the patients with severe cases of COVID-19, and our model (with high specificity) could reduce false positives—patients with a mild case of COVID-19 who were wrongly identified as having a severe case of COVID-19.

Our models were developed and validated on four different feature sets, providing the flexibility to accommodate patients with different available data. For example, if a patient does not have a chest CT scan nor a blood test, the model based on demographics and symptoms can still achieve reasonably good prediction performance (AUC 0.819, sensitivity: 0.627, specificity: 0.825). If the clinical and imaging features are available for patients, the model’s sensitivity and specificity can be improved, with potential in triaging of COVID-19 patients, e.g., prioritizing care for patients at a higher risk of mortality.

### Limitations

Our dataset consisted of 346 patients with confirmed COVID-19, with the data of 230 patients from the HSCH hospital used for training/validation and the 116 patients from the XYCH hospital used for testing. Our dataset was highly imbalanced, which could have made models overfit to the majority class. In addition, only the baseline data for patients were used in this study, therefore we could not assess how early the progression can be detected. We will be further investigating the longitudinal data and designing computational models to predict disease progression in our future work.

While we explored various configurations of NN, results were not comparable to LR, presumably due to the limited dataset and the low dimensionality of the feature vectors. In this study, we used a complex NN model (EfficientNetB7 U-Net) to extract the imaging features and tested various models for classification using the imaging features combined with tabular clinical data. Such two-stage process may simplify the classification task for these models, thereby reducing the need for another NN model for classification due to low dimensionality of features. Further exploration of NN architectures for tabular data is likely to benefit the performance of the NN model, especially if more data is available.

During training and validation, the performance of the models across cross-validation folds showed high variance due to the small number of positive cases in the validation fold. A larger dataset would improve the reliability and robustness of the models. The data also consisted of COVID-19 cases which were confirmed with RT-PCR. As such, our model is limited to differentiating severe/critical cases from mild/ordinary cases of COVID-19, and not for diagnosing COVID-19 or for differentiating COVID-19 cases from other respiratory tract infections. Further work is needed to determine the efficacy of the severity assessments, including data from asymptomatic patients.

## CONCLUSIONS

This work presents a novel method for severity assessment of diagnosed COVID-19 patients. The results indicate that clinical and imaging features can be used for automated severity assessment of COVID-19 patients. While imaging features had the strongest impact on the model’s performance, inclusion of clinical features and oversampling yielded the best performance in our study. The proposed method may have the potential to assist with triaging COVID-19 patients and prioritizing care for patients at higher risk of severe cases.

## Data Availability

The dataset is not publicly available due to patient privacy restrictions, but may be available from the corresponding author on reasonable request.

## ACKNOWLEDGMENTS

This project was supported by Natural Science Foundation of Guangdong Province (grant no.2017A030313901); Guangzhou Science, Technology and Innovation Commission (grant no.201804010239); Foundation for Young Talents in Higher Education of Guangdong Province (grant no.2019KQNCX005); and the NHMRC Centre of Research Excellence in Digital Health and the NHMRC Partnership Centre for Health System Sustainability. Dr Sidong Liu acknowledges the support of an Australian National Health and Medical Research Council grant (NHMRC Early Career Fellowship 1160760). We acknowledge Fujitsu Australia Limited for providing the computational resources for this study.

## CONTRIBUTORSHIP STATEMENT

The project was initially conceptualized and supervised by X.R.C., S.L, X.M.Q. and L.S. The patient data and imaging data were acquired by Y.Z.F., Z.Y.C. and D.R. The analysis methods were designed and implemented by J.C.Q., S.L. and H.X. The data were analyzed by J.C.Q. and S.L. The research findings were interpreted by P.K.C., Q.T.L, L.Q., X.F.L, S.B. and E.C. All authors were involved in the design of the work. The manuscript was drafted by J.C.Q., S.L and L.Q., and all authors have substantively revised it. All authors have reviewed and approved the submitted version.

